# COVID-19 pandemic dynamics in India, the SARS-CoV-2 Delta variant, and implications for vaccination

**DOI:** 10.1101/2021.06.21.21259268

**Authors:** Wan Yang, Jeffrey Shaman

## Abstract

**Background:** The COVID-19 Delta pandemic wave in India surged and declined within 3 months; cases then remained low despite the continued spread of Delta elsewhere. Here we aim to estimate key epidemiological characteristics of the Delta variant based on data from India and examine the underpinnings of its dynamics.

**Methods:** We utilize multiple datasets and model-inference methods to reconstruct COVID-19 pandemic dynamics in India during March 2020 – June 2021. We further use model estimates to retrospectively predict cases and deaths during July – mid-Oct 2021, under various vaccination and vaccine effectiveness (VE) settings to estimate the impact of vaccination and VE for non-Delta-infection recoverees.

**Findings:** We estimate that Delta escaped immunity in 34.6% (95% CI: 0 – 64.2%) of individuals with prior wildtype infection and was 57.0% (95% CI: 37.9 – 75.6%) more infectious than wildtype SARS-CoV-2. Models assuming higher VE among those with prior non-Delta infection, particularly after the 1^st^ dose, generated more accurate predictions than those assuming no such increases (best-performing VE setting: 90/95% vs. 30/67% baseline for the 1^st^/2^nd^ dose). Counterfactual modeling indicates that high vaccination coverage for 1^st^ vaccine-dose in India (∼50% by mid-Oct 2021) combined with the boosting of VE among recoverees averted around 60% of infections during July – mid-Oct 2021.

**Interpretation:** Non-pharmaceutical interventions, infection seasonality, and high coverage of 1-dose vaccination likely all contributed to pandemic dynamics in India during 2021. Given the shortage of COVID-19 vaccines globally and boosting of VE, for populations with high prior infection rates, prioritizing the first vaccine-dose may protect more people.

**Research in context:** *Evidence before this study:* We searched PubMed for studies published through Nov 3, 2021 on the Delta (B.1.617.2) SARS-CoV-2 variant that focused on three areas: 1) transmissibility [search terms: (“Delta variant” OR “B.1.617”) AND (“transmission rate” OR “growth rate” OR “secondary attack rate” OR “transmissibility”)]; 2) immune response ([search terms: (“Delta variant” OR “B.1.617”) AND (“immune evas” OR “immune escape”)]; and 3) vaccine effectiveness ([search terms: (“Delta variant” OR “B.1.617”) AND (“vaccine effectiveness” OR “vaccine efficacy” OR “vaccination”)]. Our search returned 256 papers, from which we read the abstracts and identified 54 relevant studies. Forty-two studies addressed immune evasion and/or vaccine effectiveness. Around half (n=19) of these studies measured the neutralizing ability of convalescent sera and/or vaccine sera against Delta and most reported some reduction (around 2-to 8-fold) compared to ancestral variants. The remainder (n=23) used field observations (often with a test-negative or cohort-design) and reported lower VE against infection but similar VE against hospitalization or death. Together, these laboratory and field observations consistently indicate that Delta can evade preexisting immunity. In addition, five studies reported higher B-cell and/or T-cell vaccine-induced immune response among recovered vaccinees than naïve vaccinees, suggesting potential boosting of pre-existing immunity; however, all studies were based on small samples (n = 10 to 198 individuals). Sixteen studies examined transmissibility, including 1) laboratory experiments (n=6) showing that Delta has higher affinity to the cell receptor, fuses membranes more efficiently, and/or replicates faster than other SARS-CoV-2 variants, providing biological mechanisms for its higher transmissibility; 2) field studies (n=5) showing higher rates of breakthrough infections by Delta and/or higher viral load among Delta infections than other variants; and 3) modeling/mixed studies (n=5) using genomic or case data to estimate the growth rate or reproduction number, reporting a 60-120% increase. Only one study jointly estimated the increase in transmissibility (1.3-1.7-fold, 50% CI) and immune evasion (10-50%, 50% CI); this study also reported a 27.5% (25/91) reinfection rate by Delta.

*Added value of this study:* We utilize observed pandemic dynamics and the differential vaccination coverage for two vaccine doses in India, where the Delta variant was first identified, to estimate the epidemiological properties of Delta and examine the impact of prior non-Delta infection on immune boosting at the population level. We estimate that Delta variant can escape immunity from prior wildtype infection roughly one-third of the time and is around 60% more infectious than wildtype SARS-CoV-2. In addition, our analysis suggests the large increase in population receiving their first vaccine dose (∼50% by end of Oct 2021) combined with the boosting effect of vaccination for non-Delta infection recoverees likely mitigated epidemic intensity in India during July – Oct 2021.

*Implications of all the available evidence:* Our analysis reconstructs the interplay and effects of non-pharmaceutical interventions, infection seasonality, Delta variant emergence, and vaccination on COVID-19 pandemic dynamics in India. Modeling findings support prioritizing the first vaccine dose in populations with high prior infection rates, given vaccine shortages.

## INTRODUCTION

The Delta (PANGO lineage: B.1.617.2) SARS-CoV-2 variant of concern (VOC)^1-4^ has spread quickly to over 170 countries (GISAID,^5^ as of 11/3/2021). Several lines of evidence have indicated that Delta is able to evade immunity from prior infection by preexisting variants; these include reduced neutralizing ability of convalescent sera and vaccinee sera against Delta,^6-9^ reduced vaccine effectiveness (VE) against infection,^10-13^ and reduced VE against symptomatic disease after 1-dose of vaccine (but only slight reduction for full vaccination).^14-16^ In addition, studies have found a higher secondary attack rate, growth rate, or reproduction number for Delta than prior variants including Alpha (range of the mean estimates: 60-120%).^2,17-21^ In particular, a recent study, fitting a model to mortality data in Delhi, India, estimated a 1.3-1.7-fold (50% CI) increase in transmissibility and 10-50% (50% CI) immune evasion for Delta; however, the authors noted large uncertainty in their estimates.^22^ Further, factors such as host behavioral changes and seasonal modulation of risk due to changes in environmental conditions are difficult to account for and could confound these estimates. As a result, estimates of prior immunity evasion and relative transmissibility for Delta and the contributions of these properties to the rapid spread of this variant remain uncertain.

India, where Delta was first identified, experienced an intensive pandemic wave in late March 2021. However, unlike many places seeing a prolonged Delta pandemic wave, the Delta wave in India only lasted 3 months and declined rapidly after peaking mid-May. Since June 2021 cases have remained low. A high infection rate after the Delta wave has been cited as a reason for this dramatic epidemic decline, as vaccination coverage was low at the time (4.2% fully vaccinated at the end of June 2021). However, given an estimated basic reproduction number (*R*_*0*_) of 6-7,^20^ roughly 83-86% (1 – 1/*R*_*0*_) of the population would need immunity for the Delta epidemic to subside. Assuming 10-50% immunity escape^22^ and a 25-35% infection rate prior to the Delta wave,^23^ this implies that 53-73% of India’s 1.4 billion people would have been infected by Delta within the span of 3 months, despite a national lockdown at the time.

To better understand COVID-19 pandemic dynamics in India and the epidemiological characteristics of Delta, here we utilize a model-inference method recently developed for SARS-CoV-2 VOCs. The model-inference method incorporates epidemiological, population mobility, and weather data to model SARS-CoV-2 transmission dynamics, while accounting for case under-ascertainment, impacts of non-pharmaceutical interventions (NPIs) and vaccination, infection seasonality, and new variants.^24^ Applying this method, we have jointly estimated the immune escape potential and change in transmissibility for Alpha, Beta, and Gamma, separately, using data from countries where these three VOCs were first reported.^24^ In addition, several laboratory studies have reported stronger vaccine-induced immune responses among recovered vaccinees than naïve vaccinees, suggesting potential boosting of pre-existing immunity. In India, while only 23% of the population have received two vaccine doses, 53% have received their first vaccine dose, as of the end of Oct 2021. This large discrepancy in one- and two dose coverage, combined with a likely high population infection rate, offers an opportunity to examine the boosting effect of prior non-Delta infection on vaccine-induced immunity at the population level. Therefore, in this study, we first reconstruct the pandemic dynamics in India during March 2020 – June 2021 and estimate key epidemiological characteristics of Delta. We then further use our model estimates to retrospectively predict cases and deaths during July – Oct 2021, under various vaccination and VE scenarios, and compare these simulations to observations in order to estimate the impact of vaccination and VE for those with prior non-Delta infection.

## METHODS

### Data sources and processing

We used reported COVID-19 case and mortality data to capture transmission dynamics, weather data to estimate infection seasonality, mobility data to represent concurrent NPIs, and vaccination data to account for changes in population susceptibility due to vaccination in the model-inference system. COVID-19 case and mortality data from the week of March 8, 2020 (the first week COVID-19 deaths were reported in India) to the week of Oct 17, 2021 came from the COVID-19 Data Repository of the Center for Systems Science and Engineering (CSSE) at Johns Hopkins University.^25,26^ Surface station temperature and humidity data were accessed using the “rnoaa” R package.^27^ We then aggregated these data for all weather stations in India (n = 390 stations) with measurements from Jan 2020 to Oct 2021 and calculated the average for each week of the year. Mobility data were derived from Google Community Mobility Reports;^28^ we aggregated all business-related categories (i.e., retail and recreational, transit stations, and workplaces) in all locations in India to weekly intervals. Vaccination data (1^st^ and 2^nd^ dose) were obtained from Our World in Data.^29,30^

### Model-inference system

The model-inference system was developed and described in detail in our previous study.^31^ Below we describe each component in brief.

#### Epidemic model

The epidemic model follows an SEIRSV (susceptible-exposed-infectious-recovered-susceptible-vaccination) construct per Eqn 1:

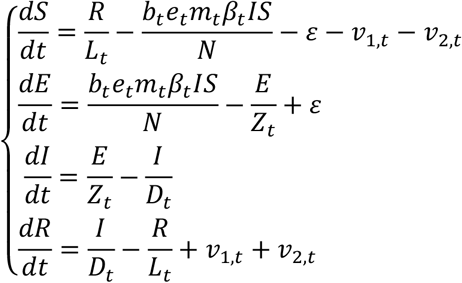

where *S, E, I, R* are the number of susceptible, exposed (but not yet infectious), infectious, and recovered/immune/deceased individuals; *N* is the population size; and *ε* is the number of travel-imported infections. In addition, the model includes the following key components:

1. Virus-specific properties, including the time-varying variant-specific transmission rate *β*_*t*_, latency period *Z*_*t*_, infectious period *D*_*t*_, and immunity period *L*_*t*_. Note the subscript, *t*, denotes time in week, as all parameters are estimated for each week as described below.
2. The impact of NPIs. Specifically, we use relative population mobility (see data above) to adjust the transmission rate via the term *m*_*t*_. To further account for potential changes in effectiveness, the model additionally includes a parameter, *e*_*t*_, to scale NPI effectiveness.
3. The impact of vaccination, via the terms *v*_*1,t*_ and *v*_*2,t*_. Specifically, *v*_*1,t*_ is the number of individuals successfully immunized after the first dose of vaccine and is computed using vaccination data and vaccine efficacy for 1^st^ dose; and *v*_*2,t*_ is the additional number of individuals successfully immunized after the second vaccine dose (i.e., excluding those successfully immunized after the first dose).
4. Infection seasonality, computed using temperature and specific humidity data as described previously (see supplemental material of Yang and Shaman^24^). The estimated relative seasonal trend, *b*_*t*_, is used to adjust the relative transmission rate at time *t*.

#### Observation model to account for under-detection and delay

Using the model-simulated number of infections occurring each day, we further computed the number of cases and deaths each week to match with the observations, as done in Yang et al.^32^ For example, for case data, we include 1) a time-lag from infectiousness to detection (i.e., an infection being diagnosed as a case) to account for delays in detection; and 2) an infection-detection rate (*r*_*t*_), i.e. the fraction of infections (including subclinical or asymptomatic infections) reported as cases, to account for under-detection. To compute the model-simulated number of new cases per week, we multiply the model-simulated number of new infections per day by the infection-detection rate, and further distribute these simulated cases in time per the distribution of time-from-infectiousness-to-detection. We then aggregate the daily lagged, simulated cases to weekly totals for model inference (see below).

#### Model inference and parameter estimation

The inference system uses the EAKF,^33^ a Bayesian statistical method, to estimate model state variables (i.e., *S, E, I, R* from Eqn 1) and parameters (i.e., *β*_*t*_, *Z*_*t*_, *D*_*t*_, *L*_*t*_, *e*_*t*_, from Eqn 1 as well as *r*_*t*_ and other parameters from the observation model). Briefly, the EAKF uses an ensemble of model realizations (*n*=500 here), each with initial parameters and variables randomly drawn from a *prior* range (see Table S1). After model initialization, the system integrates the model ensemble forward in time for a week (per Eqn 1) to compute the prior distribution for each model state variable and parameter, as well as the model-simulated number of cases and deaths for that week. The system then combines the prior estimates with the observed case and death data for the same week to compute the posterior per Bayes’ theorem.^33^ During this filtering process, the system updates the posterior distribution of all model variables and parameters for each week.

#### Estimating the immune escape potential and changes in transmissibility for Delta

To identify the most plausible combination of changes in transmissibility and level of immune evasion, per methods developed in ref^24^, we ran the model-inference, repeatedly and in turn, to test 14 major combinations of these two quantities and select the best performing run. Based on the best-performing model estimates, we then computed the variant-specific transmissibility as the product of the variant-specific transmission rate (*β*_*t*_) and infectious period (*D*_*t*_). To reduce uncertainty, we averaged transmissibility estimates over the first pandemic wave and the period when Delta is dominant, separately. We then computed the average change in transmissibility due to Delta as the ratio of the two averaged estimates (i.e., after: before the rise of Delta). To quantify immune evasion, we recorded the changes in susceptibility over time and then computed the level of immune evasion as the ratio of the total increase in susceptibility due to immune evasion during the second wave to the model-estimated population immunity at the end of the first wave.

To account for model stochasticity, we repeated the model-inference process 300 times, each with 500 model realizations and summarized the results from all 150,000 model estimates. As a sensitivity test and part of the effort to examine the impact of prior non-Delta infection on VE, we performed the analysis using 12 different VE settings (see details below).

### Model validation using independent data

To compare model estimates with independent observations not assimilated into the model-inference system, we identified three measurements of cumulative infection rates from three nationwide serology surveys in India: i) the first national serosurvey conducted during May 11 – June 4, 2020 (n = 28,000 adults 18 years or older);^34^ ii) the second national serosurvey conducted during August 18 – September 20, 2020 (n = 29,082 individuals 10 years or older);^35^ and iii) the third national serosurvey conducted during December 18, 2020 – January 6, 2021 (n = 28,598 individuals 10 years or older).^36^ To account for the delay in antibody generation, we shifted the timing of each serosurvey 14 days when comparing survey results to model-inference system estimates of cumulative infection rates in Fig 1B.

**Fig 1.**
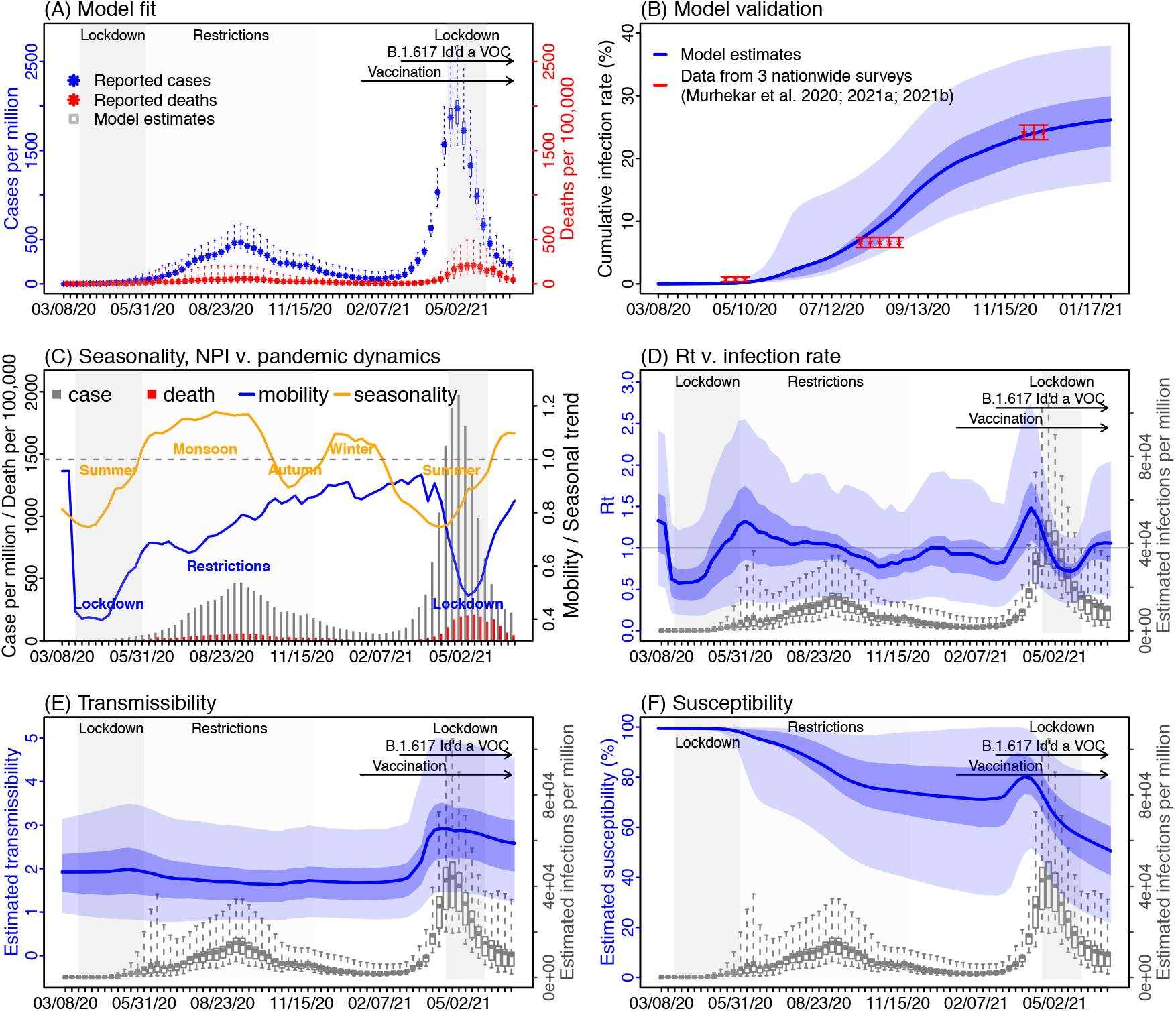
Model-inference estimates and validation. (A) Model fit. (B) Model validation. (C) Observed relative mobility and estimated disease seasonal trend, compared to case and death rates over time. Key model-inference estimates are shown for the real-time reproduction number R_t_ (D), transmissibility (E), and population susceptibility (F). Blue lines and surrounding areas show the estimated mean, 50% (dark) and 95% (light) CrIs. Boxes and whiskers show the estimated mean, 50% and 95% CrIs for weekly cases and deaths in (A) and infection rates in (D) – (F). Grey shaded areas indicate the timing of national lockdowns (darker) or local restrictions (lighter); horizontal arrows indicate the timing of variant identification and vaccination rollout. In (C), for mobility (blue line; y-axis), values below 1 (dashed horizontal line) indicate reductions due to public health interventions. For the disease seasonal trend (orange line; y-axis), values above 1 indicate weather conditions more conducive for transmission than the yearly average and *vice versa. Note that the transmissibility estimates have removed the effects of changing population susceptibility, NPIs, and disease seasonality; thus, the trends are more stable than the reproduction number (R*_*t*_; *left column) and reflect changes in variant-specific properties*.

### Estimating the impact of vaccination and prior non-Delta infection on boosting vaccine-induced immunity

We generated retrospective projections of cases and deaths from the week starting 7/4/2021 to the week starting 10/17/2021 (i.e., 16 weeks following the model-inference period), under various vaccination and VE settings. We considered four levels of VE for those recovered from non-Delta infection: 1) No boosting effect, i.e., using the same VE values as those without prior infection. Here, we set VE at fourteen days after the 1^st^ dose (VE1) to 30% and at seven days after the 2^nd^ dose (VE2) to 67%, based on data for the AstraZeneca vaccine against Delta;^15^ 2) Higher VE for the 1^st^ dose but no future boosting for the 2^nd^ dose (here, VE1 is set to 40%, 50%, or 60%, and VE2 fixed at 67%); 3) Higher VE for the 2^nd^ dose but not 1^st^ dose (here, VE1 is fixed at 30% and VE2 set to 75%, 85%, or 95%); and 4) Higher VE for both doses (here, VE1/VE2 are set to 50%/75%, 60%/80%, 70%/85%, 80%/90%, or 90%/95%). To test the impact of vaccination, in addition to projections using reported vaccination rates, we also generated counterfactual projections assuming no further vaccination during the 16-week period.

For all projections, the model was initiated using model-inference estimates made at the week of 6/27/2021, except for the infection-fatality risk (IFR). For IFR, estimates were decreasing during June 2020 (Fig S1B) and model-inference extended to the end of July 2021 showed continued decreases, likely due to improved healthcare and increased protection from prior infection or vaccination. We thus assumed that IFR would decrease linearly for the first 6 weeks of the projection period and then flatten and remain at that low IFR until the week of 10/17/2021. To account for NPIs, we used mobility data during the week of 7/4/2021 – the week of 10/17/2021. As for the model-inference runs, we repeated the projections for each scenario 300 times (each with 500 model realizations) and summarized the projections from all runs. To evaluate the projection accuracy, we computed the relative root-mean-square-error (RRMSE) and correlation between the projected and observed values for cases and deaths, respectively.

## RESULTS

### The first COVID-19 pandemic wave in India, March 2020 – January 2021

From March 2020 to January 2021 India recorded over 10 million COVID-19 cases (0.77% of its population); however, a nationwide serology survey suggested that ∼24% of its population had been infected by December 2020.^23^ Accounting for under-detection of infection (Fig S1), implemented non-pharmaceutical interventions (NPIs), seasonality, and vaccination, we used the model-inference system to reconstruct pandemic dynamics in India since March 2020 (Fig 1A). Model-estimated infection rates closely match with measurements from three nationwide serologic surveys conducted during the early, mid, and late phases of the first pandemic wave (Fig 1B). Our analysis indicates that the 2-month long national lockdown (March 24 – May 31, 2020) and the less favorable weather conditions during pre-monsoon season (i.e., March – May) likely contributed to initial low infection rates. By mid-May 2020, the model-inference system estimates that only 0.43% (95% CrI: 0.19 – 1.7%) of the population had been infected [vs. 0.73% (95% CI: 0.34%, 1.13%) among adults estimated by serosurvey^34^]. As the country lifted its lockdown in June 2020 and entered the monsoon season (June – September), when conditions are likely more favorable for transmission (Fig 1C), the first pandemic wave began. Nevertheless, continued regional restrictions during June – November 2020 and less favorable weather conditions during the autumn (October – November; see mobility and seasonal trends in Fig 1C) likely mitigated pandemic intensity. The estimated mean of the reproduction number *R*_*t*_ (i.e., average number of secondary infections per primary infection) was above 1 but less than 1.35 from June to mid-September; in addition, *R*_*t*_ dropped below 1 during October – November (Fig 1D). By the end of January 2021 when case rates reached a minimum following the first wave, the model-inference system estimates that 26.1% (95% CrI: 19.9 – 33.0%) of the population had been infected (Fig 1B).

### The second pandemic wave in India and estimated epidemiological characteristics of Delta

Infections resurged dramatically in late March 2021, largely due to the rise of the Delta variant. Despite a weeks-long second national lockdown implemented beginning April 20, 2021, India reported another 19 million cases during late March – June 2021, about twice the number reported during the previous 12 months. Accounting for under-detection (Fig S1), we estimate that 32.3% (95% CrI: 22.4 – 46.5%) of the population were infected during this 3-month period, including reinfections. This intense transmission was likely facilitated by the higher transmissibility and immune evasive capabilities of the Delta variant. Estimated transmissibility increased substantially during the second pandemic wave (Fig 1E). In addition, estimated population susceptibility increased at the start of the second pandemic wave (Fig 1F), suggesting loss of population immunity against Delta. Due to this immune escape, an estimated 50.5% (95% CrI: 21.8 – 79.0%) of the population remained susceptible at the end of June 2021, despite two large pandemic waves and rollout of mass vaccination (of note, 19% of the population had received at least 1 dose of vaccine by the end of June 2021). These findings along with the seasonal trends described above suggest that the decline of the second wave was largely due to the NPIs implemented and less favorable weather conditions during March – May, rather than high population immunity.

Combining the model-inference estimates during the first and second pandemic waves in India, we estimated that Delta was able to escape immunity among 34.6% (95% CI: 0 – 64.2%) of individuals with prior wildtype infection and was 57.0% (95% CI: 37.9 – 75.6%) more transmissible than wildtype SARS-CoV-2. Estimates are similar under different VE settings (Fig S2).

### Impact of vaccination and prior non-Delta infection on boosting vaccine-induced immunity

Despite the likely conducive conditions during the monsoon season (June – September), easing of NPIs, and relatively high susceptibility estimated at the end of June 2021, cases and deaths in India remained at relatively low levels during July – Oct 2021. Counterfactual modeling suggests that the faster rollout of vaccination during this period substantially mitigated the epidemic risk (Fig 2). Projected cases and deaths assuming no further vaccination uptake are much higher than observed; In contrast, models including the reported vaccination rates more closely match reported cases and deaths (Fig 2). Further, models assuming higher VE for non-Delta infection recoverees generated more accurate projections than those assuming no boosting effect (Fig 3). The boosting effect appears to be more pronounced for the 1^st^ vaccine dose (see, e.g., Fig 3A where larger dots, representing higher VE after the 1^st^ dose, had smaller errors). Overall, the model assuming 90%/95% VE for the 1^st^/2^nd^ dose of vaccine for non-Delta infection recoverees generated the most accurate projections. These projections estimate that vaccination rollout combined with the boosting effect averted 57% of infections during July – mid-Oct 2021.

**Fig 2.**
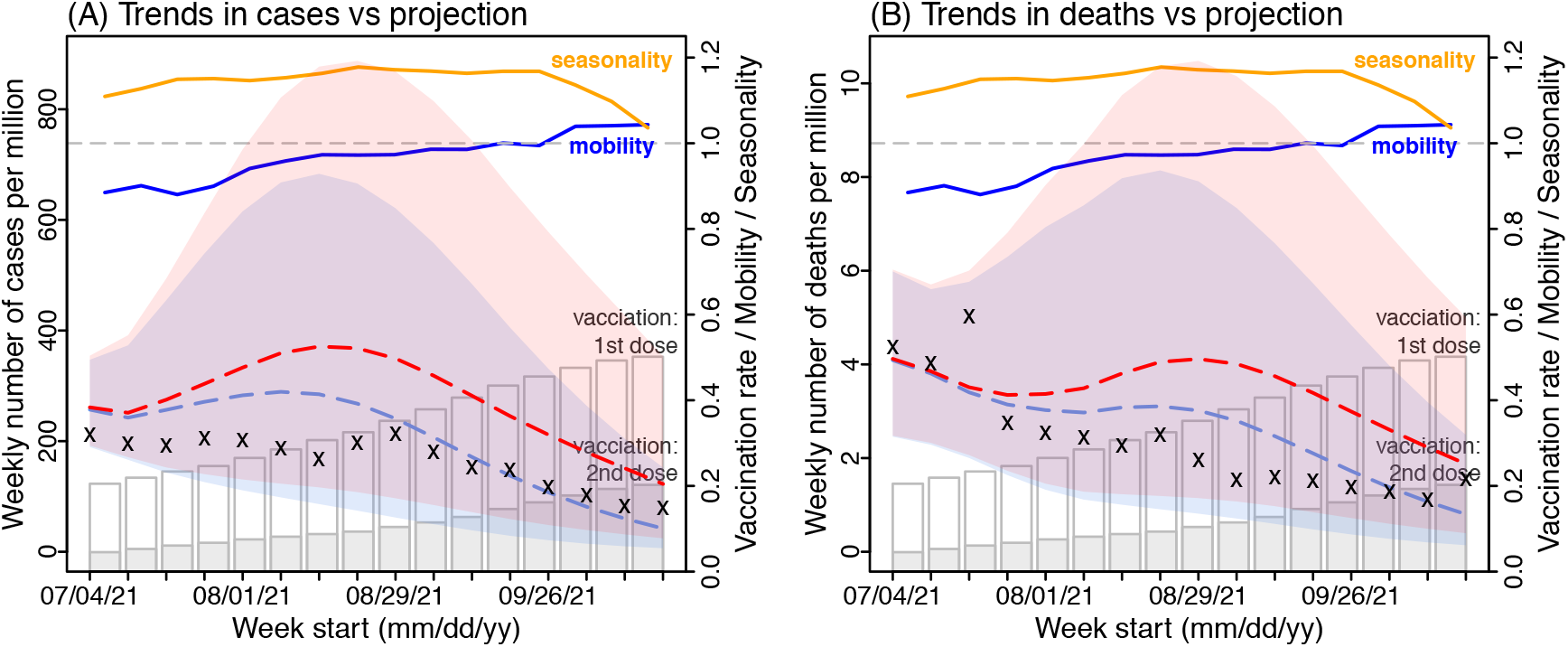
Impact of vaccination. Model projections of weekly number of reported cases (A) and reported deaths (B) for India during July – mid-Oct 2021, compared to reported data. Crosses (‘x’) show reported data (left y-axis). Red dashed lines show median counterfactual model projections assuming no further vaccination uptake during the 16-week period. Blue dashed lines show median model projections using reported vaccination rates and assuming 90%/95% vaccine effectiveness for individuals with prior non-Delta infection after the 1^st^/2^nd^ vaccine dose. Shaded areas with the same color show projected interquartile ranges. For comparison, estimated seasonality (orange lines), reported mobility (dark-blue lines), and cumulative vaccination uptake (full bar for 1^st^ dose and filled section for 2^nd^ dose) are overlaid (see right y-axis scale). All numbers are scaled per one million people.

**Fig 3.**
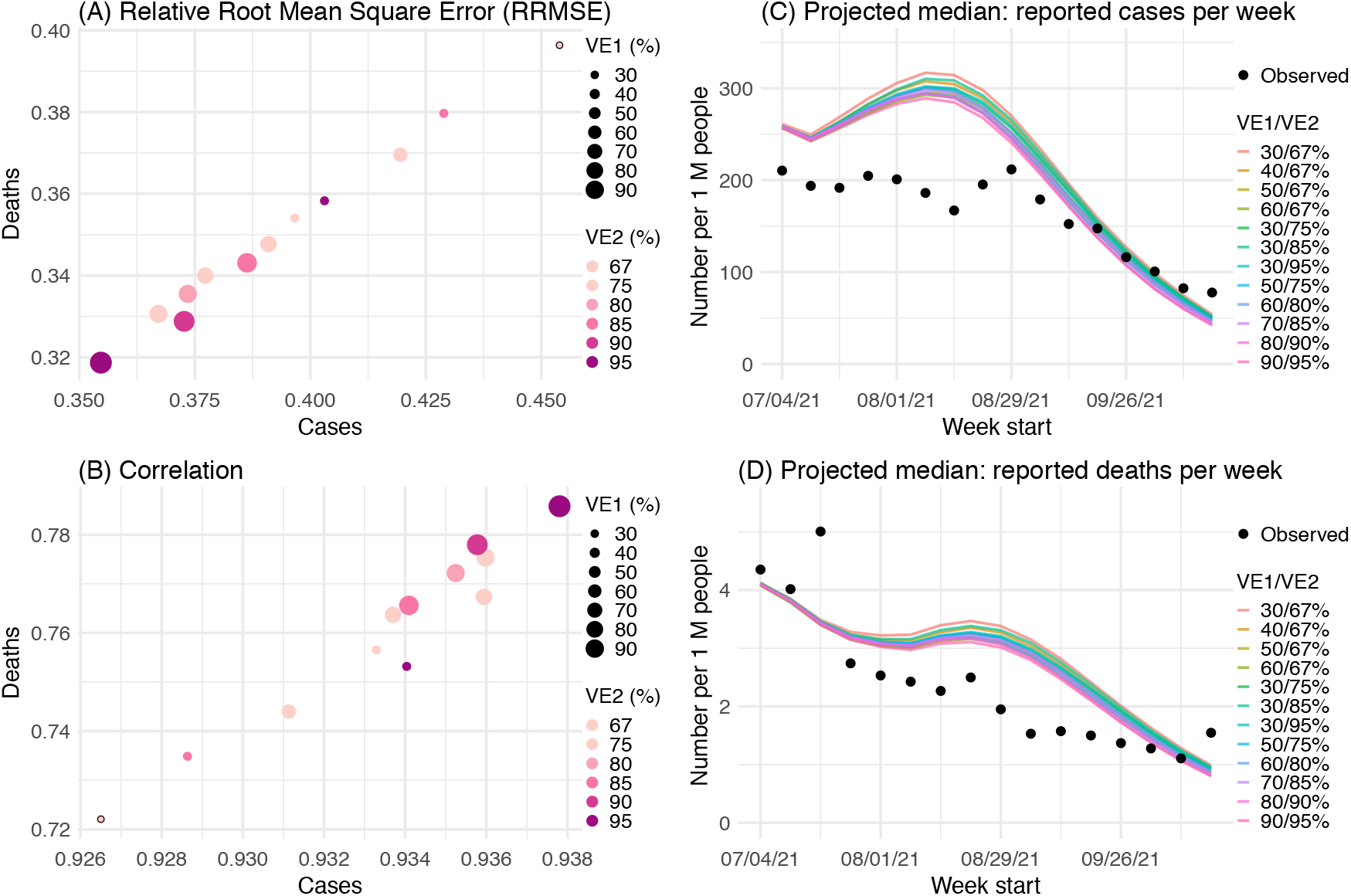
Impact of prior non-Delta infection on immune boosting. Model projections under different vaccine effectiveness (VE) settings are used to examine the most plausible VE for individuals with prior non-Delta infection, based on projection accuracy: (A) the relative root-mean-square-error (RRMSE) and (B) correlation between the projected and observed values for cases and deaths, respectively. The size of the dots represents VE for recoverees after the 1^st^ vaccine dose and the color represents VE after the 2^nd^ vaccine dose. The dots with the black circles represent the baseline VE setting (i.e. 30%/67% for the 1^st^/2^nd^ dose). For comparison, projected weekly numbers of reported cases (C) and reported deaths (D) under different VE settings are plotted along with the weekly actuals. For clarity, only median projections are shown here; see example projections including interquartile ranges in Fig 2.

## Discussion

Combining epidemiological, behavioral, and weather observational data with a comprehensive model-inference system, we estimate the Delta SARS-CoV-2 variant escaped immunity in roughly one-third of individuals with wildtype infection during the previous year and was around 60% more infectious than wildtype SARS-CoV-2. In addition, our analysis suggests the large increase in population receiving their first vaccine dose (∼50% by end of Oct 2021) combined with the boosted VE for non-Delta infection recoverees likely mitigated the epidemic intensity in India in recent months.

Previously, we have estimated the changes in transmissibility and immune escape potential for three other major SARS-CoV-2 VOCs: namely, a 46.6% (95% CI: 32.3 – 54.6%) increase in transmissibility but nominal immune escape for Alpha (i.e., B.1.1.7), a 32.4% (95% CI: 14.6 – 48.0%) increase in transmissibility and 61.3% (95% CI: 42.6 – 85.8%) immune escape for Beta (i.e., B.1.351), and a 43.3% (95% CI: 30.3 – 65.3%) increase in transmissibility and 52.5% (95% CI: 0 – 75.8%) immune escape for Gamma (i.e., P.1). Compared with Alpha, data from the UK have shown that the secondary attack rate for contacts of cases with Delta was around 1.5 times higher than Alpha (12.4% vs. 8.2%), during March 29 – May 11, 2021.^2^ In a partially immunized population, the secondary attack rate reflects the combined outcome of the transmissibility of the etiologic agent and population susceptibility to that agent. Consistent with the UK data, our estimates of the relative transmissibility and immune escape potential combine to a 44.1% (95% CI: 4.2 – 86.6%) higher secondary attack rate by Delta than Alpha [i.e., (1+57%)/(1+46.6%) × (1+34.6%) – 1 = 44.1% increase]. This higher competitiveness of Delta over Alpha explains the rapid variant displacement observed in regions previously dominated by Alpha (e.g. the UK and the US).

In addition, we estimate that 34.6% (95% CI: 0 – 64.2%) of individuals with acquired immunity from wildtype infection would be susceptible to Delta due to immune escape. This estimate is also in line with a recent study^22^ reporting a 27.5% reinfection rate during the Delta pandemic wave in Delhi, India, based on a small subset of people with repeated serology measures. In addition to immune escape from wildtype infection, studies have also reported reduced ability of sera from Beta- and Gamma-infection recoverees to neutralize Delta,^8,37^ suggesting Delta can also escape immunity conferred by those two VOCs. Such immune escape ability would also allow Delta to rapidly replace Beta and Gamma in regions previously hard-hit by those two VOCs, as has been observed in many countries in Africa and South America.^5^ More fundamentally, these findings highlight the complex, non-linear immune landscape of SARS-CoV-2 and the importance to monitor the immune escape potential of new variants against both previous and concurrent circulating variants.

Despite the successful development of multiple vaccines, shortage of supplies – particularly in resource-limited countries – remains an impediment to global mass vaccination.^38^ In response, researchers have proposed dose sparing strategies such as fractionation^39^ and 1-dose vaccination for recoverees.^40^ The latter 1-dose strategy draws on laboratory studies showing higher vaccine-induced immune response among recovered vaccinees than naïve vaccines (i.e., boosting of pre-existing immunity).^41-44^ Here, we utilized model-inference estimates and vaccination data in India to test the impact of boosting at the population level. The findings further support the effectiveness of 1-dose vaccination for recoverees. In light of continued vaccine shortages, prioritizing first-dose vaccination thus may be an effective strategy for mitigating COVID-19 burden in countries with high underlying SARS-CoV-2 infection rates.

Due to a lack of detailed epidemiological data (e.g., age-specific and subnational) and thus model simplification, our estimates have uncertainties as indicated by the large credible intervals. Nevertheless, these estimates are in line with independent data from three nationwide serology surveys conducted at three time points during the first pandemic wave in India (Fig 1B), as well as Delta-related epidemiological data from the UK^2^ and Delhi, India,^22^ as discussed above; these consistencies support the accuracy of our estimates. Unlike estimates from the contact tracing data, however, here we are able to separately quantify the changes in transmissibility and immune escape potential of the Delta variant. In addition, our analysis also suggests high 1-dose vaccine effectiveness among those with prior infection. These findings and the methods used to generate them could support better understanding of future SARS-CoV-2 variant dynamics given local prior infection rates, variant prevalence, and vaccination coverage.

## Data Availability

All data used in this study are publicly available as described in the Data sources and processing section.

https://github.com/wan-yang/covid_voc_delta

## Code availability

All source code and data necessary for the replication of our results and figures are made publicly available at https://github.com/wan-yang/covid_voc_delta.

## Acknowledgements

This study was supported by the National Institute of Allergy and Infectious Diseases (AI145883 and AI163023), the National Science Foundation Rapid Response Research Program (RAPID; DMS-2027369) and a gift from the Morris-Singer Foundation.

## Author contributions

WY designed the study, performed the analysis, and wrote the first draft; JS provided input to the analysis and contributed to the final draft.

## Supplemental Table and Figures

**Table S1.**
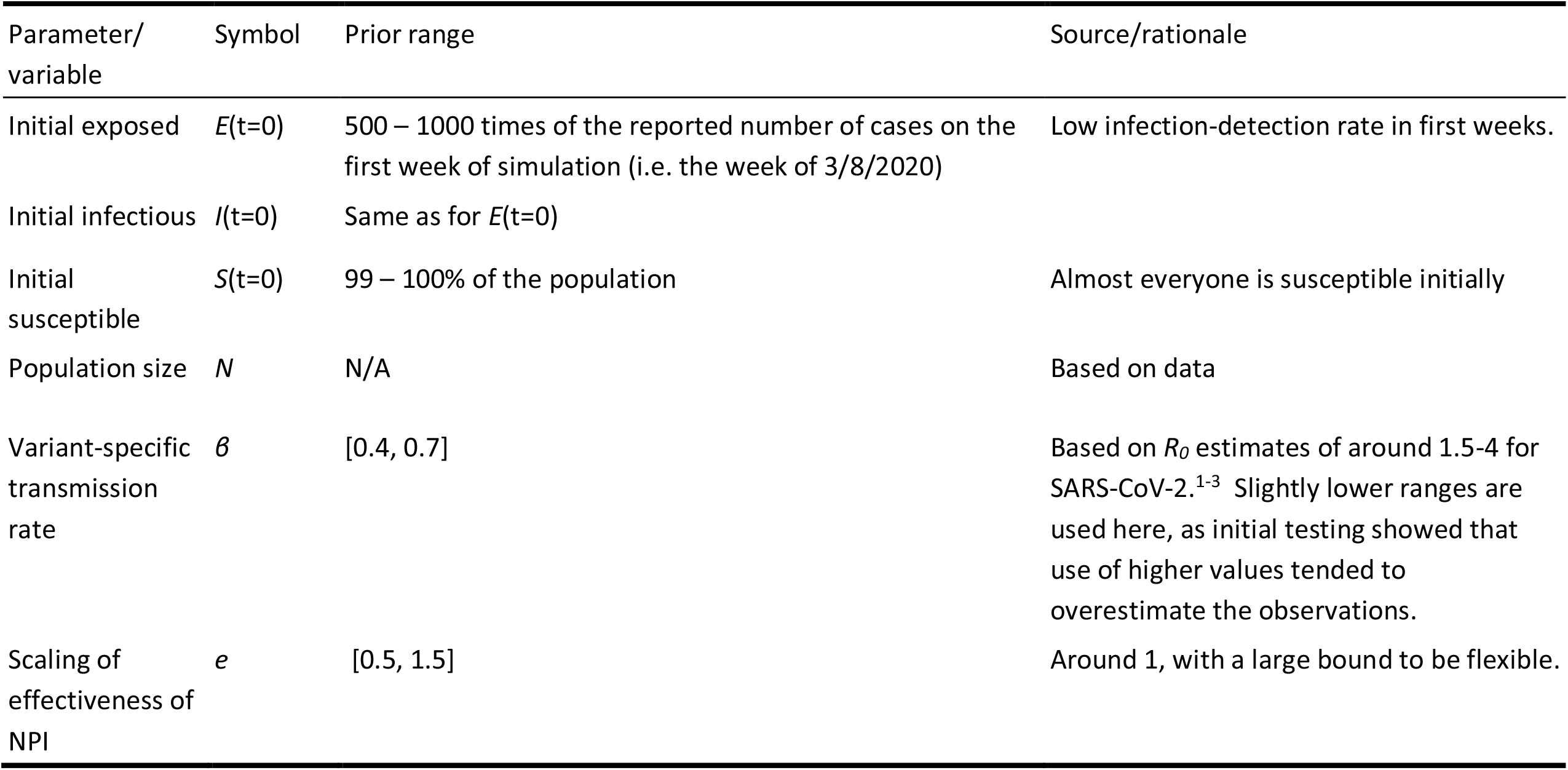
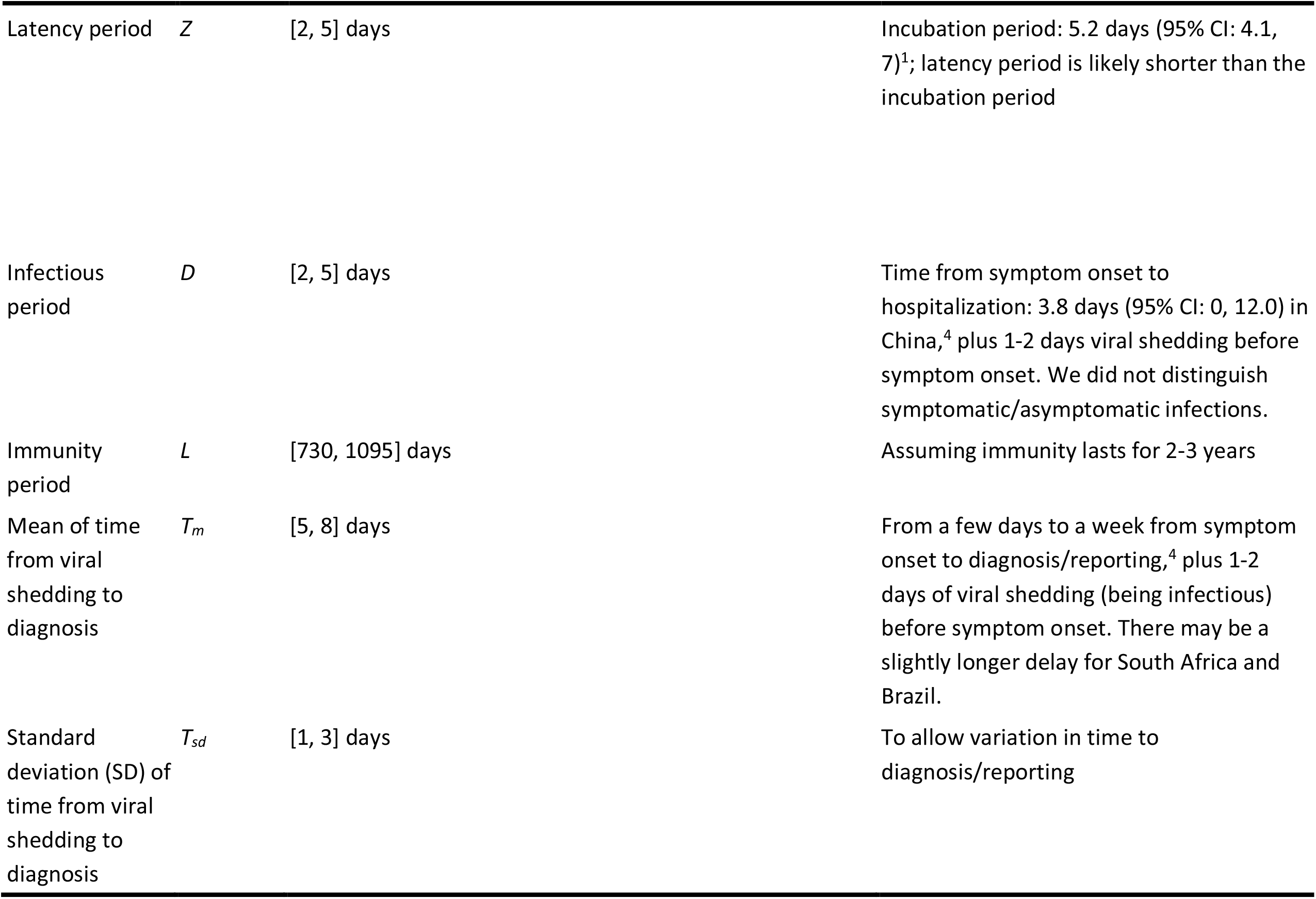
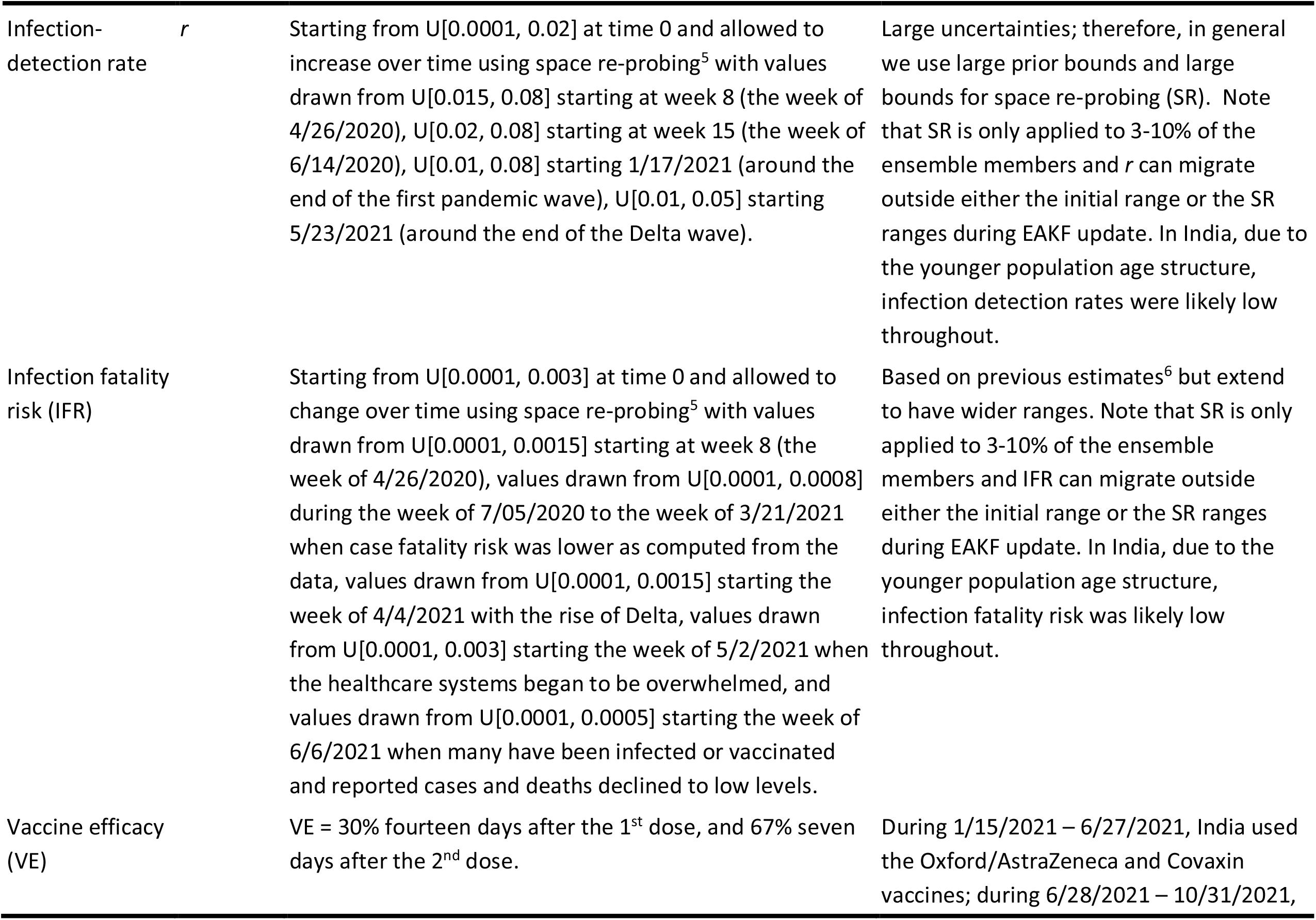
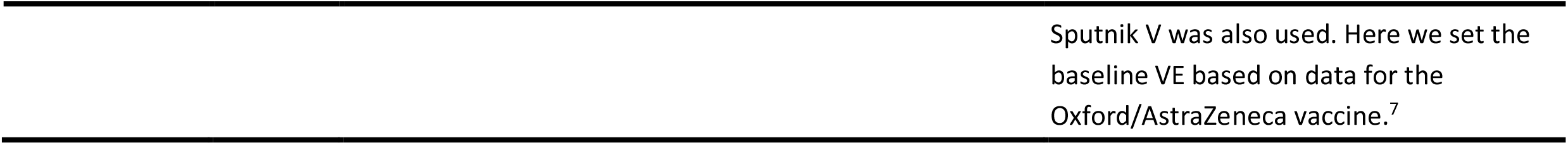
Prior ranges for the parameters used in the model-inference system.

---

**Fig S1.**
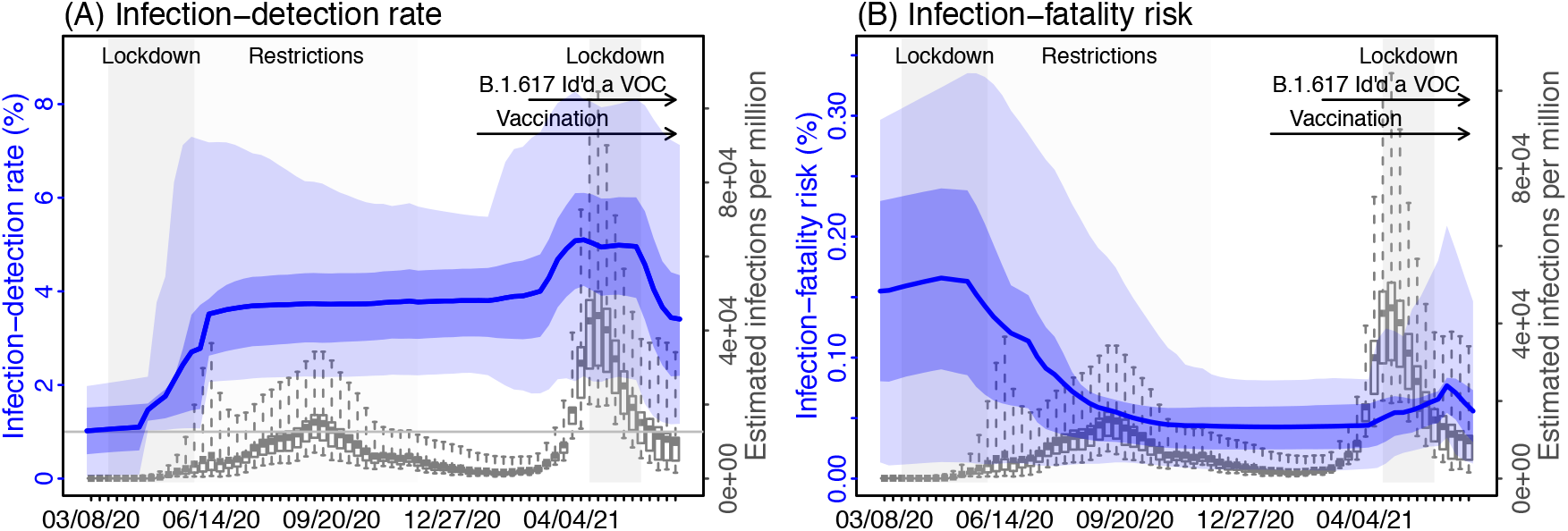
Estimated infection-detection rate (A) and infection-fatality risk (B) during each week of the study period. For comparison, estimated weekly infection rates are superimposed in each plot (right y-axis). Blue lines and surrounding areas show model estimated mean, 50% and 95% CrIs. Boxes and whiskers show model-estimated weekly infection rates (mean, 50% and 95% CrIs). Grey shaded boxes indicate the timing of lockdowns (darker) or local restrictions (lighter); horizontal arrows indicate the timing of variant identification and vaccination rollout. *Note that infection-fatality risk estimates were based on reported COVID-19 deaths and may not reflect the true values due to the likely under-reporting of COVID-19 deaths*.

**Fig S2.**
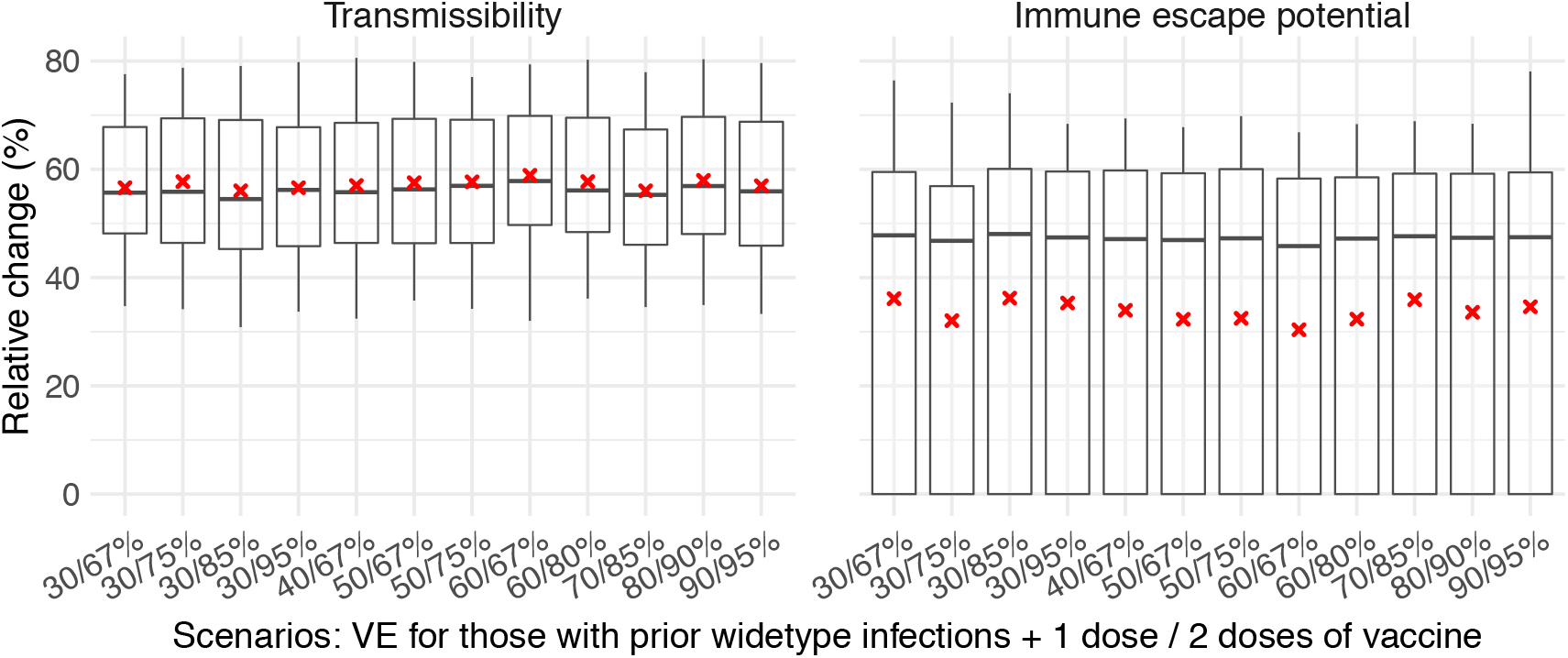
Estimated change in transmissibility and immune escape potential for the Delta variant, using different vaccine effectiveness (VE) settings for those with prior non-Delta infection. The baseline VE (i.e. 30%/67% for the 1^st^/2^nd^ dose) was used for those without any prior infection; those infected by Delta were assumed to acquire immunity against this variant after recovery (see Eqn 1 in the main text). Twelve VE settings were tested (see x-axis label for VE after 1^st^/2^nd^ dose). Model-inference was done 300 times (each with 500 ensemble members) for each VE setting. The boxplots summarize the distribution of estimates from the 300 runs: middle bar = median, edges = interquartile range, whiskers = 95% CI, red ‘x’ = mean.

## References

1. World Health Organization. Tracking SARS-CoV-2 variants. 2021. https://www.who.int/en/activities/tracking-SARS-CoV-2-variants/.

2. Public Health England. SARS-CoV-2 variants of concern and variants under investigation in England. Technical briefing 14. 2021. https://assets.publishing.service.gov.uk/government/uploads/system/uploads/attachment_data/file/991343/Variants_of_Concern_VOC_Technical_Briefing_14.pdf (accessed 6/16/2021 2021).

3. National Collaborating Centre for Infectious Diseases. Updates on COVID-19 Variants of Concern. 4/26/2021 2021. https://nccid.ca/covid-19-variants/ (accessed 5/3/2021 2021).

4. Centers for Disease Control and Prevention. SARS-CoV-2 Variant Classifications and Definitions. 2021. https://www.cdc.gov/coronavirus/2019-ncov/variants/variant-info.html (accessed 6/17/2021 2021).

5. Global Initiative on Sharing All Influenza Data (GISAID). Tracking of Variants. 6/16/2021 2021. https://www.gisaid.org/hcov19-variants/ (accessed 6/16/2021 2021).

6. Mlcochova P, Kemp SA, Dhar MS, et al. SARS-CoV-2 B.1.617.2 Delta variant replication and immune evasion. Nature 2021.

7. Wall EC, Wu M, Harvey R, et al. Neutralising antibody activity against SARS-CoV-2 VOCs B.1.617.2 and B.1.351 by BNT162b2 vaccination. Lancet 2021.

8. Liu C, Ginn HM, Dejnirattisai W, et al. Reduced neutralization of SARS-CoV-2 B.1.617 by vaccine and convalescent serum. Cell 2021; 184(16): 4220–+.

9. Arora P, Sidarovich A, Krüger N, et al. B.1.617.2 enters and fuses lung cells with increased efficiency and evades antibodies induced by infection and vaccination. Cell Reports 2021; 37(2): 109825.

10. Chemaitelly H, Tang P, Hasan MR, et al. Waning of BNT162b2 Vaccine Protection against SARS-CoV-2 Infection in Qatar. N Engl J Med 2021.

11. Goldberg Y, Mandel M, Bar-On YM, et al. Waning Immunity after the BNT162b2 Vaccine in Israel. N Engl J Med 2021.

12. Puranik A, Lenehan PJ, Silvert E, et al. Comparison of two highly-effective mRNA vaccines for COVID-19 during periods of Alpha and Delta variant prevalence. medRxiv 2021: 2021.08.06.21261707.

13. Elliott P, Haw D, Wang H, et al. Exponential growth, high prevalence of SARS-CoV-2, and vaccine effectiveness associated with the Delta variant. Science 2021: eabl9551.

14. Bernal JL, Andrews N, Gower C, et al. Effectiveness of COVID-19 vaccines against the B.1.617.2 variant. medRxiv 2021: 2021.05.22.21257658.

15. Bernal JL, Andrews N, Gower C, et al. Effectiveness of Covid-19 Vaccines against the B.1.617.2 (Delta) Variant. New England Journal of Medicine 2021; 385(7): 585–94.

16. Seppala E, Veneti L, Starrfelt J, et al. Vaccine effectiveness against infection with the Delta (B.1.617.2) variant, Norway, April to August 2021. Eurosurveillance 2021; 26(35).

17. Allen H, Vusirikala A, Flannagan J, et al. Household transmission of COVID-19 cases associated with SARS-CoV-2 delta variant (B.1.617.2): national case-control study. The Lancet regional health Europe 2021: 100252.

18. Challen R, Dyson L, Overton CE, et al. Early epidemiological signatures of novel SARS-CoV-2 variants: establishment of B.1.617.2 in England. medRxiv 2021: 2021.06.05.21258365.

19. Earnest R, Uddin R, Matluk N, et al. Comparative transmissibility of SARS-CoV-2 variants Delta and Alpha in New England, USA. medRxiv 2021.

20. Vohringer HS, Sanderson T, Sinnott M, et al. Genomic reconstruction of the SARS-CoV-2 epidemic in England. Nature 2021.

21. Campbell F, Archer B, Laurenson-Schafer H, et al. Increased transmissibility and global spread of SARS-CoV-2 variants of concern as at June 2021. Eurosurveillance 2021; 26(24).

22. Dhar MS, Marwal R, Vs R, et al. Genomic characterization and epidemiology of an emerging SARS-CoV-2 variant in Delhi, India. Science 2021: eabj9932.

23. Murhekar MV, Bhatnagar T, Thangaraj JWV, et al. SARS-CoV-2 seroprevalence among the general population and healthcare workers in India, December 2020–January 2021. Int J Infect Dis 2021; 108: 145–55.

24. Yang W, Shaman J. Development of a model-inference system for estimating epidemiological characteristics of SARS-CoV-2 variants of concern. Nature Communications 2021; 12: 5573.

25. COVID-19 Data Repository by the Center for Systems Science and Engineering (CSSE) at Johns Hopkins University. 2021. https://github.com/CSSEGISandData/COVID-19.

26. Dong E, Du H, Gardner L. An interactive web-based dashboard to track COVID-19 in real time. Lancet Infect Dis 2020; 20(5): 533–4.

27. Chamberlain S, Anderson B, Salmon M, et al. Package ‘rnoaa’. 2021.

28. Google Inc. Community Mobility Reports. 2020. https://www.google.com/covid19/mobility/.

29. Data on COVID-19 (coronavirus) vaccinations by Our World in Data. 2020. https://github.com/owid/covid-19-data/tree/master/public/data/vaccinations.

30. Mathieu E, Ritchie H, Ortiz-Ospina E, et al. A global database of COVID-19 vaccinations. Nat Hum Behav 2021; 5(7): 947–53.

31. Yang W, Shaman J. Epidemiological characteristics of three SARS-CoV-2 variants of concern and implications for future COVID-19 pandemic outcomes. medRxiv 2021: 2021.05.19.21257476.

32. Yang W, Kandula S, Huynh M, et al. Estimating the infection-fatality risk of SARS-CoV-2 in New York City during the spring 2020 pandemic wave: a model-based analysis. The Lancet Infectious diseases 2021; 21(2): 203–12.

33. Anderson JL. An ensemble adjustment Kalman filter for data assimilation. Mon Weather Rev 2001; 129(12): 2884–903.

34. Murhekar MV, Bhatnagar T, Selvaraju S, et al. Prevalence of SARS-CoV-2 infection in India: Findings from the national serosurvey, May-June 2020. Indian J Med Res 2020; 152(1): 48–57.

35. Murhekar MV, Bhatnagar T, Selvaraju S, et al. SARS-CoV-2 antibody seroprevalence in India, August-September, 2020: findings from the second nationwide household serosurvey. The Lancet Global Health 2021; 9(3): e257–e66.

36. Murhekar MV, Bhatnagar T, Thangaraj JWV, et al. SARS-CoV-2 seroprevalence among the general population and healthcare workers in India, December 2020 - January 2021. Int J Infect Dis 2021; 108: 145–55.

37. de Oliveira T, Lessells R. Update on Delta and other variants in South Africa and other world. 2021. https://www.krisp.org.za/manuscripts/DeltaGammaSummary_NGS-SA_6JulV2.pdf.

38. Padma TV. COVID vaccines to reach poorest countries in 2023 — despite recent pledges. 7/5/2021 2021. https://www.nature.com/articles/d41586-021-01762-w (accessed 11/11/2021 2021).

39. Cowling BJ, Lim WW, Cobey S. Fractionation of COVID-19 vaccine doses could extend limited supplies and reduce mortality. Nature Medicine 2021; 27(8): 1321–3.

40. Sasikala M, Shashidhar J, Deepika G, et al. Immunological memory and neutralizing activity to a single dose of COVID-19 vaccine in previously infected individuals. International Journal of Infectious Diseases 2021; 108: 183–6.

41. Andreano E, Paciello I, Piccini G, et al. Hybrid immunity improves B cells and antibodies against SARS-CoV-2 variants. Nature 2021.

42. Lucas C, Vogels CBF, Yildirim I, et al. Impact of circulating SARS-CoV-2 variants on mRNA vaccine-induced immunity. Nature 2021.

43. Sokal A, Barba-Spaeth G, Fernández I, et al. mRNA vaccination of naive and COVID-19-recovered individuals elicits potent memory B cells that recognize SARS-CoV-2 variants. Immunity 2021.

44. Sapkal GN, Yadav PD, Sahay RR, et al. Neutralization of Delta variant with sera of Covishield™ vaccinees and COVID-19-recovered vaccinated individuals. J Travel Med 2021; 28(7).

## References

1. Li Q, Guan X, Wu P, et al. Early Transmission Dynamics in Wuhan, China, of Novel Coronavirus–Infected Pneumonia. New Engl J Med 2020.

2. Wu JT, Leung K, Leung GM. Nowcasting and forecasting the potential domestic and international spread of the 2019-nCoV outbreak originating in Wuhan, China: a modelling study. Lancet 2020; 395(10225): 689–97.

3. Li R, Pei S, Chen B, et al. Substantial undocumented infection facilitates the rapid dissemination of novel coronavirus (SARS-CoV-2). Science 2020; 368(6490): 489–93.

4. Zhang J, Litvinova M, Wang W, et al. Evolving epidemiology and transmission dynamics of coronavirus disease 2019 outside Hubei province, China: a descriptive and modelling study. The Lancet Infectious diseases 2020; 20(7): 793–802.

5. Yang W, Shaman J. A simple modification for improving inference of non-linear dynamical systems. arXiv 2014: 1403.6804.

6. Verity R, Okell LC, Dorigatti I, et al. Estimates of the severity of coronavirus disease 2019: a model-based analysis. The Lancet Infectious diseases 2020; 20(6): 669–77.

7. Bernal JL, Andrews N, Gower C, et al. Effectiveness of Covid-19 Vaccines against the B.1.617.2 (Delta) Variant. New England Journal of Medicine 2021; 385(7): 585–94.

